# Sociodemographic disparities in the use of GLP-1 receptor agonists and SGLT-2 inhibitors among US adults with type 2 diabetes: NHANES 2005-March 2020

**DOI:** 10.1101/2023.03.30.23287965

**Authors:** Benjamin G. Mittman, Phuc Le, Julia Y. Payne, Gina Ayers, Michael B. Rothberg

## Abstract

**Aim:** Type 2 Diabetes (T2D) is a major cause of morbidity and mortality. Glucagon-like peptide-1 receptor agonists (GLP-1 RAs) and sodium-glucose cotransporter-2 inhibitors (SGLT-2i) are highly effective but underutilized. We assessed racial/ethnic and other sociodemographic disparities in GLP-1/SGLT-2 use among US adults with T2D.

**Materials and Methods:** We conducted a retrospective analysis using nationally representative data from National Health and Nutrition Examination Survey 2005-March 2020. Participants were adults with T2D taking ≥1 diabetes medication, excluding pregnant women and adults with probable T1D. We performed univariate analyses to examine characteristics of patients using GLP-1/SGLT-2 and multivariable logistic regression to assess disparities in GLP-1/SGLT-2 use after adjusting for other patient factors.

**Results:** Among 4,585 T2D patients (representing >18 million US adults) taking ≥1 medication, GLP-1/SGLT-2 usage increased from 1.4% in 2005-2006 to 13.3% in 2017-2020. In univariate analyses, patients using GLP-1/SGLT-2 vs. other T2D drugs were more likely to be white than nonwhite (72% vs. 60%, p = .001), but in multivariable analysis there was no significant difference in GLP-1/SGLT-2 use for nonwhite vs. white patients (aOR = 0.84, 95% CI [0.61, 1.16]). GLP-1/SGLT-2 use was higher for patients who completed some college (aOR = 1.83, 95% CI [1.06, 3.15]) or above (aOR = 2.06, 95% CI [1.28, 3.32]) vs. high school or less, and for those with an income-poverty ratio ≥4 vs. <2 (aOR = 2.11, 95% CI [1.30, 3.42]).

**Conclusions:** Use of GLP-1/SGLT-2 drugs increased over time but remained low in March 2020. Higher education and income, but not race/ethnicity, were associated with GLP-1/SGLT-2 use.

## Introduction

Type 2 Diabetes (T2D) is a major cause of morbidity and mortality in adults, affecting approximately 37 million individuals (11% of all adults) with an overall mortality rate of 31 per 100,000 people in the United States (US).^1^ Pharmacological agents, including biguanides, sulfonylureas, meglitinides, and thiazolidinediones, have been successfully used to improve glycemic control for decades.^2,3^ Two newer antihyperglycemic drug classes, glucagon-like peptide-1 receptor agonists (GLP-1 RAs) and sodium-glucose cotransporter-2 inhibitors (SGLT-2i), first approved to manage T2D in 2005 and 2013 respectively, also control hyperglycemia but have additional clinical benefits.^4^ They support weight loss in patients with comorbid obesity,^5^ lower the risk of cardiovascular events,^6^ slow the progression of kidney disease,^7^ and reduce all-cause mortality.^8^

However, these new drugs cost substantially more than older drugs, posing a potential barrier to access. Newer medications, including those belonging to GLP-1/SGLT-2 classes, can cost as much as 1,730% more than older, less effective medications.^9^ Moreover, the ability to afford even standard medications varies greatly based on race/ethnicity, socioeconomic status, and insurance coverage, particularly for patients with comorbid conditions that incur annual expenses.^10–12^

Multiple studies have addressed concerns about high costs and other barriers to access by examining patterns of GLP-1/SGLT-2 medication use among adults with T2D. Many of these studies have revealed disparities in the uptake of these drugs in the US. Sociodemographic disparities, especially along race/ethnicity, have been documented in multiple subpopulations including Medicare patients,^13,14^ veterans,^15,16^ older adults,^13,17,18^ and patients with comorbid cardiovascular disease (CVD)^13,18^ and chronic kidney disease (CKD).^13^ Low socioeconomic status has also been associated with worse access to these drugs, independent of insurance status, potentially due to the impact of high co-pays or deductibles.^19,20^ Disparities in the use of GLP-1/SGLT-2 drugs within racial/ethnic minority groups and other marginalized populations are particularly detrimental because these groups face a higher burden of chronic conditions for which these medications have added benefit, especially atherosclerotic CVD and CKD.^19^

A majority of studies have examined disparities within a specific subpopulation. It remains an open question as to whether these sociodemographic disparities affect the US population as a whole. One study examined disparities in the receipt of these drugs in a national sample, but its assessment was limited by a short time period and separate analysis of GLP-1 and SGLT-2 use, consistent with most other studies.^21^ The lack of analysis of use of either GLP-1 or SGLT-2 is important because for most patients receipt of either drug would offer substantial benefit due to significant overlap in their indications and therapeutic effects. To our knowledge, there are no studies assessing sociodemographic disparities in a nationally representative US sample across the full time period for which either GLP-1 and SGLT-2 drugs have been available. Our objective was to assess racial/ethnic and other sociodemographic disparities in the receipt of either GLP-1 or SGLT-2 drugs, after adjusting for other patient factors, in a nationally representative sample of US adults with a diagnosis of T2D.

## Materials and Methods

### Data Source

We conducted a retrospective analysis using continuous National Health and Nutrition Examination Survey (NHANES) data from 2005-March 2020. NHANES is a nationally representative survey in the US, administered by the National Center for Health Statistics (NHCS) of the Centers for Disease Control and Prevention, which combines interviews, physical examinations, and laboratory tests to assess the health and nutritional status of noninstitutionalized adults and children in the United States.^22^ Our study population included participants aged ≥18 years who had ever been told they had diabetes by their doctor, had an HbA1C >6.4%, or had a fasting plasma glucose >125 mg/dL and who took at least one diabetes medication. We excluded pregnant women and patients with probable type 1 diabetes (defined as age <20 years or age at diagnosis of diabetes <20 years and use of insulin as their only medication).

### Outcomes and Predictors

The primary outcome of interest was the use of GLP-1 RA/SGLT-2i, with or without other diabetes drugs. The main predictors were key patient sociodemographic characteristics: race/ethnicity (categorized as white or nonwhite); insurance (categorized as none, private, or government); and socioeconomic status, represented by education level (categorized as less than high school, some college, or college or above) and family income-poverty ratio (categorized as <2, 2-4, or ≥4). We adjusted for the following patient factors, including additional sociodemographic characteristics and cardiovascular/renal comorbidities common in T2D: age, sex, BMI, HbA1C %, smoking status (categorized as never, former, or current), and common T2D comorbidities (hypertension, hypertriglyceridemia, obesity, coronary artery disease (CAD), angina, heart attack, heart failure, stroke, and chronic kidney disease (CKD); each categorized as yes or no).

### Statistical Analyses

We first estimated the prevalence of using GLP-1 RAs/SGLT-2i with or without other diabetes drugs (GLP-1/SGLT-2) aggregately for the whole study period, and for each individual survey cycle. For the whole study population, we compared characteristics of patients who received GLP-1/SGLT-2 to those receiving only other diabetes medications using independent samples t-tests for continuous variables and chi-square tests or analyses of variance (ANOVAs) for categorical variables.

These univariate analyses were used to assess individual relationships with GLP-1/SGLT-2 use and identify potentially important predictors; characteristics with a p-value <0.2 were subsequently included as predictors in a multivariable logistic regression model performed to investigate the relationships between key patient characteristics and GLP-1/SGLT-2 use. Multiple imputation using chained equations was employed for missing observations among predictor variables. The multivariable analysis was performed on the imputed data to assess the relationships between race/ethnicity, insurance, education, and income and use of GLP-1/SGLT-2, after adjusting for other patient characteristics. For all analyses, 95% confidence intervals (CIs) were computed around point estimates. We considered a p-value of <0.05 as statistically significant. We used Stata 16.1 for all analyses and accounted for the complex survey design of NHANES.

## Results

The final sample included 4,585 T2D patients who received ≥1 diabetes medication, representing more than 18 million adults. The prevalence of use of GLP-1/SGLT-2 increased significantly from 1.4% in 2005-2006 to 13.3% in 2017-March 2020; overall, almost 7% of treated patients used them. Use of GLP-1 alone remained low from 2005-2006 to 2011-2012, then increased in 2013-2014 and plateaued, while SGLT-2 use increased consistently from 2013-2014 to 2017-March 2020 (Figure).

**Figure:**
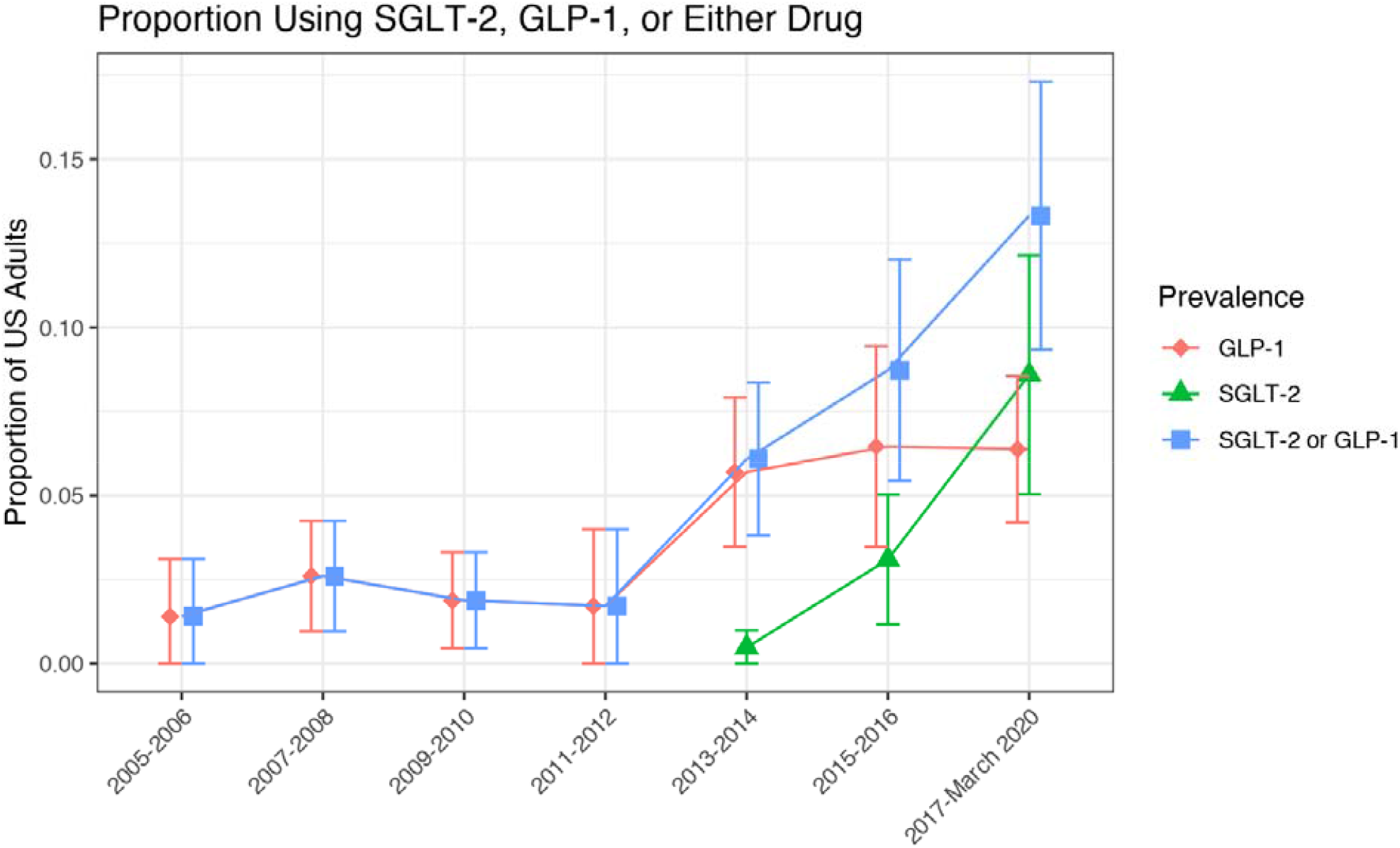
The prevalence of using GLP-1, SGLT-2, or both in US adults with type 2 diabetes, NHANES 2005-March 2020. GLP-1, glucagon-like peptide-1 receptor agonists; SGLT-2, sodium-glucose cotransporter-2 inhibitors

Patients using GLP-1/SGLT-2 were significantly younger (57.0 vs. 61.7 years, p < 0.001); more likely to be White (72% vs. 60%, p = .001) and have private insurance (72% vs. 55%, p = .014); had a higher mean BMI (35.7 vs. 33.1 kg/m^2^, p <0.001), education (29% vs. 20% completed college or above, p < .001) and income (48% vs. 27% had a IPR >= 4, p < .001); and lower prevalence of hypertension (32% vs. 50%, p < .001) and chronic kidney disease (15% vs. 24%, p = .028) than those using other drug classes. There was no significant difference between the two groups in sex, mean HbA1c, smoking, or other comorbidities (all p > .16; Table 1).

**Table 1:** Characteristics of US adults with type 2 diabetes who received pharmacotherapy, NHANES 2005-March 2020

|  | **GLP-1/SGLT-2** | **Other diabetes drugs** | **P value** |
| --- | --- | --- | --- |
|  | (n=220) | (n=4,361) |  |
|  | Mean/Percentage (95% CI) | Mean/Percentage (95% CI) |  |
| **Age, mean** | 57 (54.9-59.2) | 61.7 (61.1-62.3) | <0.001 |
| **BMI (kg/m<sup>2</sup>), mean** | 35.7 (34.6-36.8) | 33.1 (32.7-33.4) | <0.001 |
| **HbA1C (%), mean** | 7.4 (7.1-7.7) | 7.4 (7.4-7.5) | 0.758 |
| **Sex** |  |  | 0.161 |
| Female | 41 (31-51) | 48 (46-51) |  |
| Male | 59 (49-69) | 52 (49-54) |  |
| **Race** |  |  | 0.001 |
| White | 72 (65-78) | 60 (56-63) |  |
| Nonwhite | 28 (22-35) | 40 (37-44) |  |
| **Education** |  |  | <0.001 |
| Less than high school | 9 (5-14) | 24 (22-26) |  |
| High school & Some College | 61 (50-72) | 56 (54-58) |  |
| College and above | 29 (20-39) | 20 (17-22) |  |
| **Family income to poverty ratio** |  |  | <0.001 |
| <2 | 22 (16-28) | 43 (40-46) |  |
| 2 - <4 | 30 (20-40) | 30 (27-32) |  |
| >=4 | 48 (37-59) | 27 (25-30) |  |
| **Insurance** |  |  | 0.014 |
| None | 2 (0-5) | 8 (7-9) |  |
| Private | 72 (63-82) | 55 (53-58) |  |
| Government | 25 (17-34) | 37 (34-39) |  |
| **Smoking status** |  |  | 0.875 |
| Never | 52 (43-62) | 50 (48-52) |  |
| Former | 35 (25-44) | 36 (34-38) |  |
| Current | 13 (7-19) | 14 (12-15) |  |
| **Comorbidities** |  |  |  |
| Hypertension | 32 (24-40) | 50 (48-52) | <0.001 |
| Hypertriglyceridemia | 25 (16-33) | 24 (22-25) | 0.832 |
| Obesity | 78 (70-86) | 63 (61-66) | 0.002 |
| Coronary artery disease | 17 (11-24) | 13 (12-15) | 0.192 |
| Angina | 9 (2-15) | 8 (7-10) | 0.890 |
| Heart attack | 14 (6-22) | 11 (10-13) | 0.504 |
| Heart failure | 11 (5-17) | 10 (9-12) | 0.876 |
| Stroke | 7 (2-12) | 10 (8-11) | 0.375 |
| Chronic kidney disease | 15 (8-21) | 24 (22-26) | 0.028 |

Note: BMI, body mass index; CI, confidence interval; GLP-1, glucagon-like peptide-1 receptor agonists; HbA1c, hemoglobin A1c; NHANES, National Health and Nutrition Examination Survey; SGLT-2, sodium-glucose cotransporter-2 inhibitors

**Table 2:** Association of patient characteristics and use of GLP-1/SGLT-2 in US adults with type 2 diabetes who received pharmacotherapy, NHANES 2005-March 2020

| Patient characteristics | Univariate Analysis |  | Multivariable Analysis |  |
| --- | --- | --- | --- | --- |
|  | OR (95% CI) | P value | aOR (95% CI) | P value |
| **Age** | 0.97 (0.96-0.98) | <0.001 | 0.99 (0.97-1) | 0.064 |
| **BMI, kg/m<sup>2</sup>** | 1.04 (1.02-1.06) | <0.001 | 1.04 (1.02-1.07) | <0.001 |
| **HbA1C, %** | 0.98 (0.88-1.1) |  | - |  |
| **Sex** |  |  |  |  |
| Female | reference |  | reference |  |
| Male | 1.34 (0.89-2.03) | 0.161 | 1.22 (0.78-1.91) | 0.382 |
| **Race** |  |  |  |  |
| White | reference |  | reference |  |
| Nonwhite | 0.58 (0.42-0.8) | 0.001 | 0.84 (0.61-1.16) | 0.291 |
| **Education** |  |  |  |  |
| Less than high school | reference |  | reference |  |
| High school & Some College | 2.92 (1.63-5.22) | <0.001 | 1.83 (1.06-3.15) | 0.030 |
| College and above | 4.00 (2.37-6.76) | <0.001 | 2.06 (1.28-3.32) | 0.003 |
| **Family income to poverty ratio** |  |  |  |  |
| <2 | reference |  | reference |  |
| 2 - <4 | 1.96 (1.25-3.07) | 0.004 | 1.44 (0.91-2.28) | 0.122 |
| >=4 | 3.39 (2.16-5.32) | <0.001 | 2.11 (1.3-3.42) | 0.003 |
| **Insurance** |  |  |  |  |
| None | reference |  | reference |  |
| Private | 4.56 (1.18-17.61) | 0.028 | 3.04 (0.76-12.14) | 0.115 |
| Government | 2.42 (0.68-8.64) | 0.173 | 2.63 (0.72-9.6) | 0.141 |
| **Smoking status** |  |  |  |  |
| Never | reference |  | reference |  |
| Former | 0.92 (0.61-1.38) | 0.678 | - |  |
| Current | 0.91 (0.53-1.55) | 0.72 | - |  |
| **Comorbidities** |  |  |  |  |
| Hypertension | 0.48 (0.33-0.71) | <0.001 | 0.6 (0.41-0.88) | 0.009 |
| Hypertriglyceridemia | 1.05 (0.65-1.69) | 0.832 | - |  |
| Obesity | 2.07 (1.32-3.24) | 0.002 | - |  |
| Coronary artery disease | 1.35 (0.86-2.14) | 0.192 | 1.58 (0.95-2.62) | 0.077 |
| Angina | 1.06 (0.45-2.49) | 0.890 | - |  |
| Heart attack | 1.26 (0.64-2.48) | 0.504 | - |  |
| Heart failure | 1.05 (0.55-2.01) | 0.876 | - |  |
| Stroke | 0.7 (0.31-1.55) | 0.375 | - |  |
| Chronic kidney disease | 0.55 (0.33-0.94) | 0.028 | 0.68 (0.35-1.32) | 0.251 |

Note: BMI, body mass index; CI, confidence interval; GLP-1, glucagon-like peptide-1 receptor agonists; HbA1c, hemoglobin A1c; NHANES, National Health and Nutrition Examination Survey; SGLT-2, sodium-glucose cotransporter-2 inhibitors

The multivariable logistic regression model included race/ethnicity, education level, income-poverty level, and insurance status as predictors. The model also included the following adjustment variables, which had a p-value <0.2 in univariate analysis: age, sex, BMI, and three comorbidities (hypertension, CAD, and chronic kidney disease). In the multivariable analysis, the odds of using GLP-1/SGLT-2 drugs was not significantly different for nonwhite vs. White patients (aOR = 0.84, 95% CI [0.61, 1.16]), or for patients with private (aOR = 3.04, 95% CI [0.76, 12.14]) or government insurance (aOR = 2.63, 95% CI [0.72, 9.60]) compared with no insurance. However, the odds of using GLP-1/SGLT-2 drugs was significantly greater for patients who completed some college (aOR = 1.83, 95% CI [1.06, 3.15]) or college education or above (aOR = 2.06, 95% CI [1.28, 3.32]) compared with patients who completed up to high school. The odds of GLP-1/SGLT-2 use was also higher for those with an income-poverty ratio ≥4 compared with those who had a ratio <2 (aOR = 2.11, 95% CI [1.30, 3.42]).

## Discussion

In this study of US adults with T2D who received pharmacotherapy, we did not find evidence of statistically significant insurance disparities in the use of highly effective GLP-1/SGLT-2 drugs, although point estimates suggested large effects may exist. Similarly, point estimates showing differences in GLP-1/SGLT-2 use by race/ethnicity were not statistically significant, in contrast to many prior studies performed with nonnational samples.^14,19,20,23^ Although univariate analyses showed disparities along these two sociodemographic factors, these effects were attenuated and not significant in multivariable analysis, after adjusting for other patients factors. Nevertheless, we did find that higher education and income were each independently associated with an increased likelihood of taking these drugs.

We also observed that the use of GLP-1/SGLT-2 drugs increased over time from 2005-2006 to 2017-March 2020, at which cycle the prevalence was approximately 13%, representing substantial underutilization. Growing evidence supporting the effectiveness and low side effect profiles of these drugs, along with high rates of comorbidity of their major indications (e.g., ASCVD, CKD, obesity) with T2D, indicate that these drugs should be incorporated into the treatment regimens of a majority (>80%) of patients with T2D.^24,25^ However, we have shown that as of three years ago, most eligible T2D patients did not receive newer highly effective GLP-1/SGLT-2 drugs. This is consistent with many other studies demonstrating substantial underutilization.^20,23,25–28^ In 2020, the revised American Diabetes Association (ADA) guidelines for standards of care for T2D patients strengthened recommendations for these drugs as first-line treatments for T2D patients with comorbid renal or cardiovascular disease, due to growing evidence of their benefits beyond glycemic control.^29^ Thus, it will be important to continue assessing equitable access to these drugs as uptake increases along with stronger guideline recommendations.

These findings make several important contributions to our understanding of the prevalence and distribution of sociodemographic disparities in GLP-1/SGLT-2 use across the US. First, income and education were associated with these drugs but race/ethnicity and insurance coverage were not. The high cost of these drugs likely poses a substantial barrier to access, regardless of insurance or any racial biases. For example, one study showed that for patients with Medicare—whose coverage of GLP-1/SGLT-2 drugs ranges from 3%-95% depending on the specific plan and drug—median estimated annual out-of-pocket costs are $1211 for ertugliflozin (an SGLT-2i) and $2447 for liraglutide (a GLP-1 RA).^30^ Another study examining drug prices under Medicare found that monthly list prices for GLP-1/SGLT-2 drugs range from $434-$935, compared with $3-$11 for metformin, sulfonylureas, and thiazolidinediones.^31^ Thus, many patients with low socioeconomic status may not be able to afford GLP-1/SGLT-2 drugs even if they have insurance, due to the dramatically higher out-of-pocket and co-pay costs compared to older first-line medications.

An additional barrier to access may be the lack of awareness of the effectiveness of these drugs among patients with low socioeconomic status or among physicians whom those patients tend to visit. More research is needed to understand how lower education and income act as barriers to GLP-1/SGLT-2 use. This research will be especially important as overall utilization of these drugs climbs. The recently published consensus report from the American Diabetes Association and European Association for the Study of Diabetes recommending GLP-1/SGLT-2 use among patients with T2D and comorbid renal/cardiovascular disease highlights growing support for wider use of these medications.^32^ However, without a complete understanding of modifiable barriers to access (e.g., costs, awareness), disparities are at risk of increasing as utilization continues to grow. Research on the roles of education and income will enable more targeted efforts to mitigate underutilization among disadvantaged and minority groups. There have been recent calls to develop frameworks and solutions, incorporating principles of health equity, to increase uptake and expand access to GLP-1/SGLT-2 drugs.^33^ However, disparities cannot be adequately addressed without a complete understanding of sociodemographic barriers,^34^ including those which have been demonstrated in the current study.

Key strengths of our study were the inclusion of a representative sample of general US adults with T2D and assessment of disparities using 15 years of continuous data, starting from the introduction of GLP-1/SGLT-2 drugs. However, our study had important limitations related to the data source, including exclusion of residents of nursing homes or other long-term care facilities who might have different patterns of use, potential inadvertent inclusion of patients with type 1 diabetes, and the small number of patients who received GLP-1/SGLT-2 in our sample, which limited our ability to detect racial/ethnic or insurance disparities after adjusting for other patient characteristics. We were also unable to determine specific reasons why many patients were not taking these drugs.

In summary, we did not find robust evidence of disparities in GLP-1/SGLT-2 use among US adults with T2D based on race/ethnicity or insurance, contrary to prior studies. However, we found evidence of substantial underutilization of GLP-1/SGLT-2 drugs, as well as disparities based on education and income. Further research, including with larger population-based samples, is needed to investigate reasons for underutilization and fully assess the effects of income and education on access to newer, better diabetes drugs.

## Data Availability

All data produced are available online at https://www.cdc.gov/nchs/nhanes/.

## Conflict of interest

none

## Notes

### Competing Interest Statement

The authors have declared no competing interest.

### Funding Statement

This study was funded by NIH grants 5T32GM007250-45 and 5TL1TR002549-04.

### Author Declarations

The study used ONLY openly available human data that were originally located at: https://www.cdc.gov/nchs/nhanes/.

